# CHEST DRAIN AEROSOL GENERATION IN COVID-19 AND EMISSION REDUCTION USING A SIMPLE ANTI-VIRAL FILTER

**DOI:** 10.1101/2020.07.13.20152264

**Authors:** Clodagh Duffy, Andrew C. Kidd, Sarah Francis, Selina Tsim, Laura McNaughton, Katie Ferguson, Jenny Ferguson, Gary Rodgers, Claire McGroarty, Robin Sayer, Kevin G. Blyth

## Abstract

**Introduction:** The COVID-19 pandemic has been characterised by significant in-hospital virus transmission and deaths among healthcare workers. Sources of in-hospital transmission are not fully understood, with special precautions currently reserved for procedures previously shown to generate aerosols (particles <5 microns). Pleural procedures are not currently considered AGPs, reflecting a lack of data in this area.

**Methods:** An underwater seal chest drain bottle (R54500, Rocket Medical UK) was set up inside a 60-litre plastic box and connected via an airtight conduit to a medical air supply. A multichannel particle counter (TSI Aerotrak 9310 Aerosol Monitor) was placed inside the box, allowing measurement of particle count/cubic foot (pc/ft^3^) within six channel sizes: 0.3-0.5, 0.5-1, 1-3, 3-5, 5-10 and >10 microns. Stabilised particle counts at 1, 3 and 5 L/min were compared by Wilcoxon signed rank test; p-values were Bonferroni-adjusted. Measurements were repeated with a simple anti-viral filter, designed using repurposed materials by the study team, attached to the drain bottle. The pressure within the bottle was measured to assess any effect of the filter on bottle function.

**Results:** Aerosol emissions increased with increasing air flow, with the largest increase observed in smaller particles (0.3-3 microns). Concentration of the smallest particles (0.3-0.5 microns) increased from background levels by 700, 1400 and 2500 pc/ft^3^ at 1, 3 and 5 L/min, respectively. However, dispersion of particles of all sizes was effectively prevented by use of the viral filter at all flow rates. Use of the filter was associated with a maximum pressure rise of 0.3 cm H_2_O after 24hours of flow at 5 L/min, suggesting minimal impact on drain function.

**Conclusion:** A bubbling chest drain is a source of aerosolised particles, but emission can be prevented using a simple anti-viral filter. These data should be considered when designing measures to reduce in-hospital spread of SARS-CoV-2.

## INTRODUCTION

Previous coronavirus epidemics were characterised by high infection rates in healthcare workers (HCWs) and ‘super-spreading’ events within hospitals. Despite implementation of World Health Organisation (WHO) guidance designed to reduce in-hospital spread, [1] nosocomial and HCW infection have remained prominent features of the current COVID-19 pandemic, with 43.5% of UK HCWs becoming seropositive over a 1-month period in one recent study. [2] Better understanding of in-hospital infection sources is therefore urgently needed. Pleural procedures are not currently considered aerosol-generating by the WHO [1], and special precautions to mitigate against viral transmission and/or to protect HCWs are not currently recommended. However, this is based on the absence of any prior data regarding the aerosol generating potential of a chest drain (where an aerosol is defined as a particle smaller than 5 microns) rather than a robust understanding of the level of risk involved. Recent studies report detection of severe acute respiratory syndrome coronavirus 2 (SARS-CoV-2) in COVID-19-associated pleural effusion [3,4] and ‘super-spreading’ events linked to chest drain use. The latter includes a cohort of 25 Chinese patients (including 12 HCWs) infected from a single index case who underwent elective lobectomy with undetected SARS-CoV-2 infection. [5] This has prompted expert bodies, including the British Thoracic Society (BTS) [6] and the American Association for the Surgery of Trauma (AAST) [7] to recommend risk mitigation of some form until the level of risk involved is more clearly understood. The objectives of this study were to determine a) whether a bubbling chest drain is aerosol generating and b) the efficacy of a simple anti-viral filter.

## METHODS

The experimental set-up used is shown in Figure 1. An underwater seal chest drain bottle (R54500, Rocket Medical UK, Ltd) was set up according to the manufacturer’s instructions and placed inside a sealable 60-litre plastic box. The drain tubing (Ref R54502) was attached to a medical air cylinder via an airtight conduit in the wall of the box. A multichannel particle counter (TSI Aerotrak 9310 Aerosol Monitor) was placed inside the box, allowing measurement of particle count/cubic foot (pc/ft^3^) within six channel sizes: 0.3-0.5, 0.5-1, 1-3, 3-5, 5-10 and >10 microns. Stabilised particle counts at different flow rates (without and with the filter attached) were compared by Wilcoxon signed rank test (R version 4.0.2 (Vienna, Austria)); p-values were adjusted for multiple comparisons by the Bonferroni method. For filter comparisons, relative differences, normalised to without-filter readings, were assessed. Ethical approval was not required.

**Figure 1.**
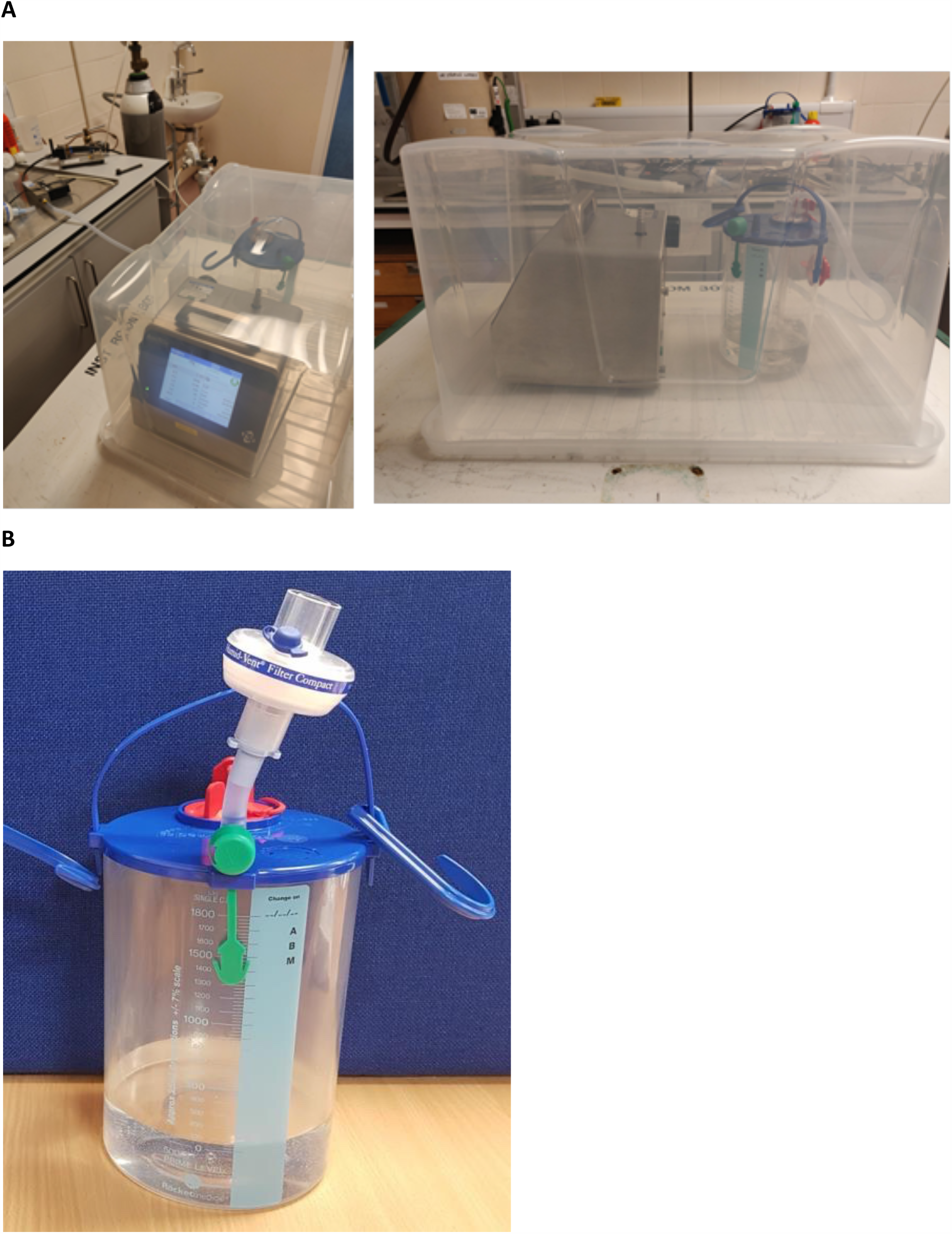
**A:** Experimental setup including the Aerotrak 9310 (to the left of the images) and the underwater seal chest drain bottle (Rocket R54500, to the right of the images) inside the sealed 60 litre plastic box. Standard chest drain tubing (R54502) has been used to connect the chest drain bottle to a medical air cylinder. **B:** The assembled COVID-19 anti-viral filter, comprised of a Heat and Moisture Exchange (HME) filter (Teleflex Humid-Vent Filter Compact, 19402T), attached via a 5 cm section of standard chest drain tubing (Rocket Medical R54502) and the proximal adapter from a size 8 endotracheal tube (Portex 100/199/080). Detailed instructions for use are provided in the online supplementary appendix.

### Viral Filter

A COVID-19 anti-viral filter device was designed by the study team and manufactured using repurposed equipment (see Figure 1, and online supplement for detailed Instructions for Use). The completed assembly was comprised of a Heat and Moisture Exchange (HME) filter normally used on a ventilator circuit (Teleflex Humid-Vent Filter Compact, 19402T), a 5 cm section of standard chest drain tubing (Rocket Medical R54502) and the proximal adapter from a size 8 endotracheal tube (Portex 100/199/080). The safe functioning of the filter was assessed within a Failure Mode and Effects Analysis (FMEA) framework (BS EN ISO 14971:2012: Application of risk management to medical devices). This included demonstration that over 24 hours of continuous flow at 5L/minute, the pressure within the bottle rose by no more than 0.3 cm H_2_O, suggesting the filter has minimal impact on drain function.

### Aerosol Measurement

Particle concentrations in the air surrounding the chest drain were initially sampled without the filter attached. Baseline conditions were first sampled for 20 minutes. Medical air was then pumped through the circuit for 20 minutes at 1 L/minute, before being switched off, allowing baseline conditions to re-stabilise over a further 20 minutes. After each 60-minute experiment the box was opened, measurements were recorded, and the unit resealed. The experiment was repeated using flow rates of 3 and 5 L/minute, and then all three experiments were repeated with the filter attached (see Figure 1(b)).

## RESULTS

Particle concentrations measured in each channel at 1, 3 and 5 L/minute, without the filter attached are shown in Figure 2. Particle emissions increased with increasing air flow, with the largest increase observed in the smaller particles (0.3-3 microns). Concentration of the smallest particles (0.3-0.5 microns) increased from background levels by 700, 1400 and 2500 pc/ft^3^ at 1, 3 and 5 L/min, respectively (see Figure 2). Particle counts (pc/ft^3^) significantly increased (based on Bonferroni adjusted p<0.00086) at all flow rates and in all channel sizes, except at 1L/minute in the 5-10 micron channel (adjusted p-value not significant at 0.02).

**Figure 2.**
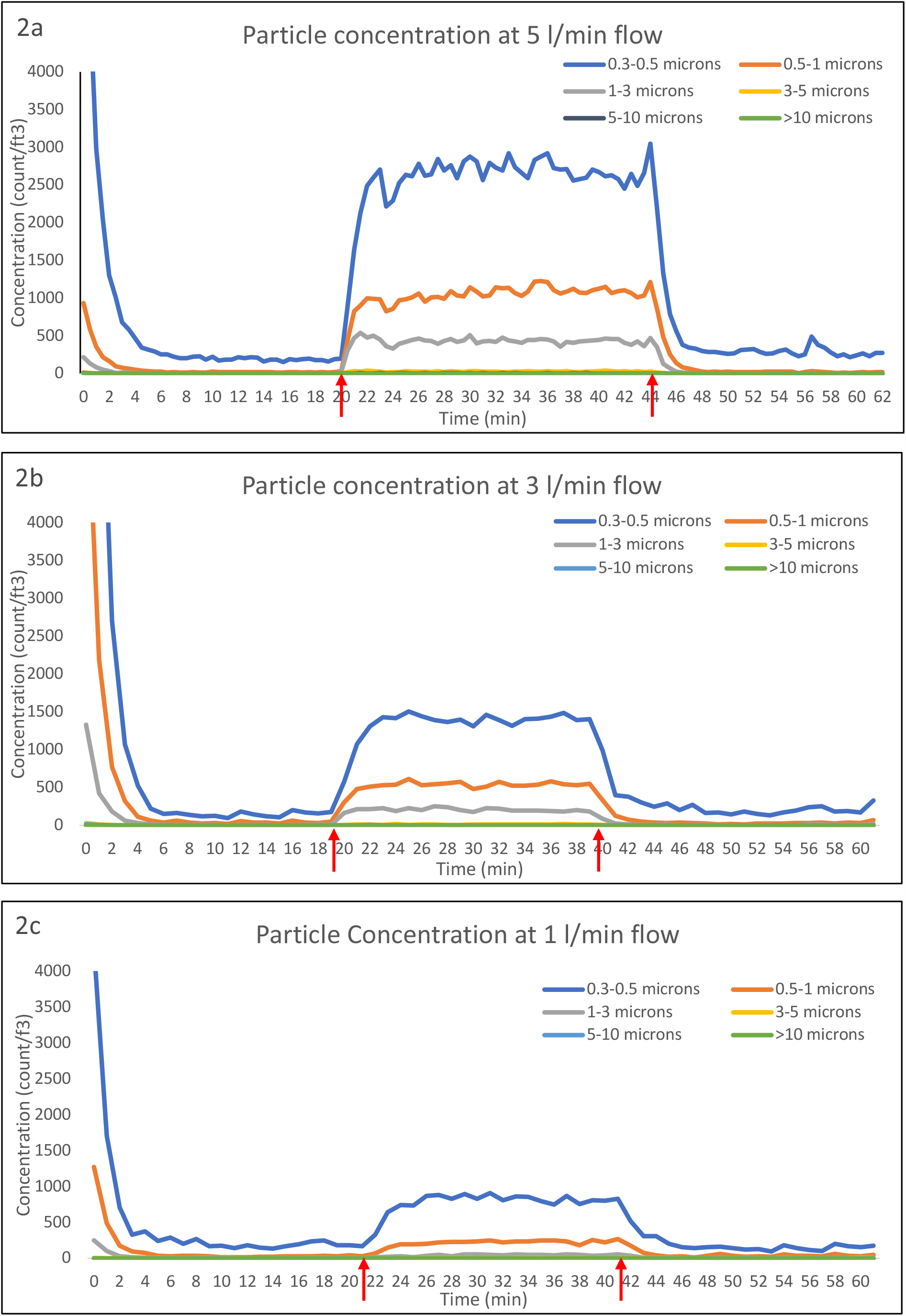
Particle concentrations, as measured by the TSI Aerotrak 9310 Aerosol Monitor, while air was bubbled through a standard underwater seal chest drain (Rocket R54500) **without a filter** at flow rates of a) 1 b) 3 and c) 5 L/minute. The red arrows on each graph show when the air flow was turned on and then turned off again.

Particle concentrations measured in each channel at 1, 3 and 5 L/minute, with the filter attached are shown in Figure 3. Particle counts (pc/ft^3^) were notably lower in all channels compared to without-filter measurements. Counts of the smallest particles significantly reduced between baseline and initiation of air flow (based on Bonferroni adjusted p<0.00086) at 1 L/minute (0.3-0.5 micron only), 3 and 5 L/minute (0.3-0.5, 0.5-1, 1-3 micron channels), likely reflecting dilution of background aerosols by filtered air. With the filter added, normalised particle count differences were significantly lower (p<0.00086) in the smaller channels (0.3-0.5, 0.5-1, 1-3, 3-5 micron) at all flow rates, compared to without-filter values.

**Figure 3.**
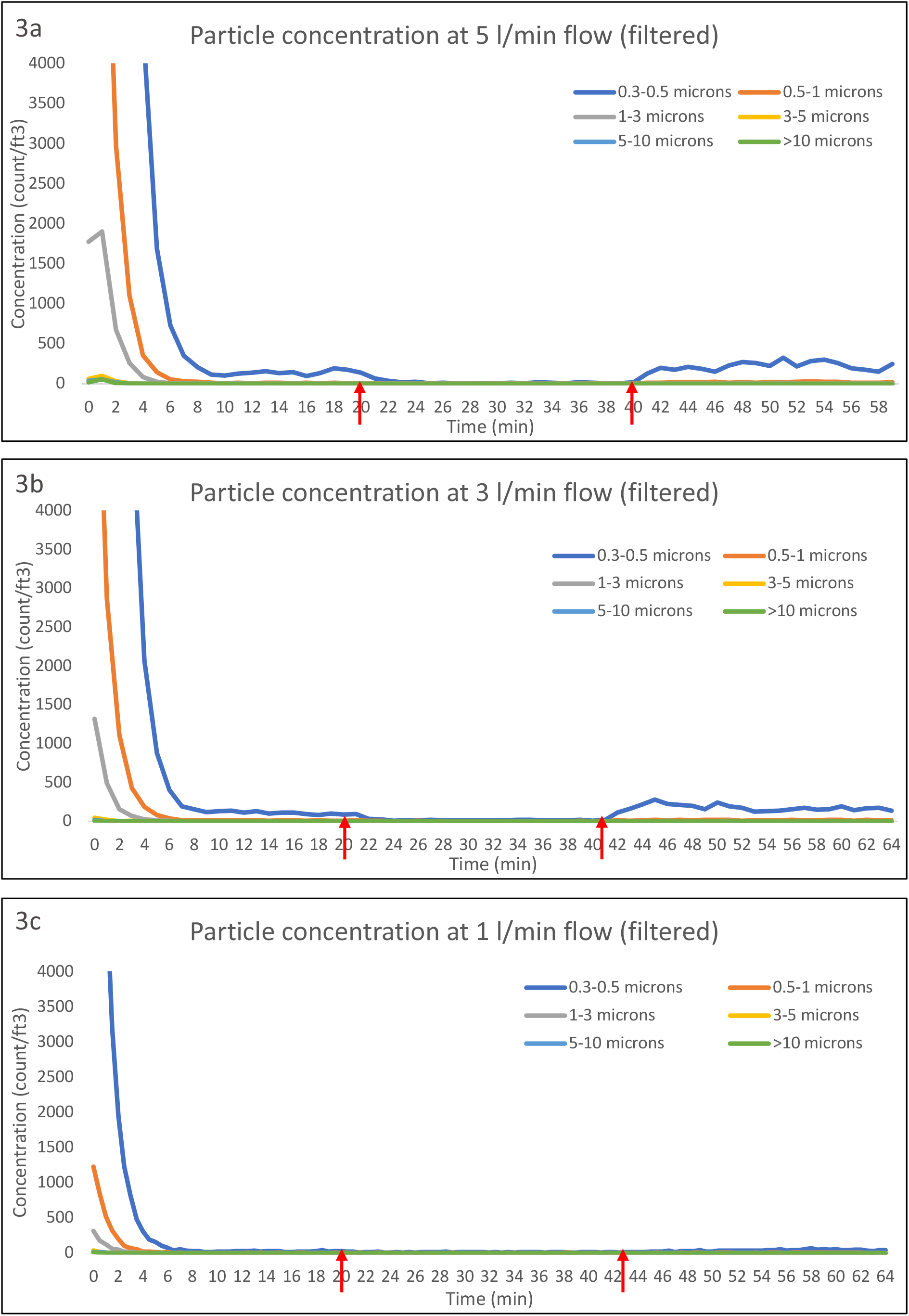
Particle concentrations, as measured by the TSI Aerotrak 9310 Aerosol Monitor, while air was bubbled through a standard underwater seal chest drain (Rocket R54500) **with a filter** at flow rates of a) 1 b) 3 and c) 5 litres per minute. The red arrows on each graph show when the air flow was turned on and then turned off again.

## DISCUSSION

The data reported here indicate that particles in the aerosol range (<5 microns) are generated by a bubbling chest drain at continuous flow rates of at least 1 L/minute. Aerosol emissions increased with increasing air flow, simulating a high-volume air-leak, as might be expected following thoracic surgery, in mechanically ventilated patients or patients with a large spontaneous alveolar-pleural fistula complicating bullous lung disease. These data were recorded as part of a comprehensive risk assessment process, which demonstrated no significant limitation of air flow through the chest drain circuit with the filter in-situ.

Aerosols are typically generated by air moving across the surface of a liquid, with increasing air forces generating smaller particles. [1] This is consistent with the observations reported here in that unfiltered emissions of the smallest particles (0.3-0.5 microns) increased progressively from 1 to 3 to 5 L/minute. Our findings are also concordant with a recent study reported by Akhtar et al, in which a similar anti-viral filter was evaluated and produced a qualitative reduction in droplet emissions. However, droplet size, and therefore aerosolization potential were not examined. [8] These data support risk mitigation in patients with suspected or proven COVID-19, as recommended by the British Thoracic Society (BTS) [6] and the American Association for the Surgery of Trauma (AAST). [7] However, the absolute risk involved remains uncertain. Pleural effusion and pneumothorax appear uncommon complications of COVID-19 (occurring in ∼5% and ∼1% of cases, respectively [6][8]) and an aerosol-generating chest drain can clearly only be an infection risk if SARS-CoV-2 is a) present in any effusion drained (which may be of minimal volume in patients with pneumothorax and major air-leaks) and b) remains viable long enough to be aerosolised. Lescure [3] and Mei [4] have recently reported cases of SARS-CoV-2 positive effusions, but in these cases air-leak was not reported as a significant component, and larger studies are needed to define the prevalence and risks of unfiltered air-leaks in COVID19 more clearly. In a recent post-mortem series, Schaller et al reported that 50% of patients with fatal COVID-19 had associated pleural effusion, of which 50% were PCR positive. [9] The risk of nosocomial transmission via an unfiltered air leak may therefore be highest in patients with the most advanced disease in the event of a secondary pneumothorax complicating positive pressure ventilation. With regard to viability over time, SARS-CoV-2 has been shown to remain viable in aerosols for several hours and on surfaces for several days, [10] so could probably persist in a chest drain bottle for sufficiently long to be an infection risk.

The experiments reported here were carried out in a controlled environment with minimal background environmental disturbances. It is therefore unknown how normal background activities (e.g. staff proximity, the opening and closing of doors), which can affect the rate of aerosols resuspension on surfaces, would affect concentrations in a clinical setting. Given that our data were recorded at flow rates of 1, 3 and 5 L/minute it should also be acknowledged that the particle emission profile of smaller volume air-leaks cannot be directly concluded from our data. Nevertheless, it appears clear that a bubbling chest drain is a source of aerosolised particles, and that dispersion can be prevented using a simple anti-viral filter. These data should be considered when designing measures to reduce in-hospital spread of SARS-CoV-2.

## Data Availability

The data is available upon reasonable request to the corresponding author.

## ACKNOWLEDEMENTS

CD had full access to all of the data in the study, contributed substantially to the study design, data analysis and interpretation, and wrote a first draft of part of the manuscript

AK contributed substantially to the study design, and interpretation, and wrote a first draft of part of the manuscript

SF and RS had full access to all of the data in the study, contributed substantially to the study design, data analysis and interpretation, and the writing of the manuscript

ST, KF, JF, LM, GR, CM contributed substantially to the study design, data analysis and interpretation, and the writing of the manuscript

KGB conceived the study, had access to the data and takes responsibility for the integrity and the accuracy of the data analysis, had the primary role in writing and submitting the manuscript and acts as guarantor.

